# Epidemiology, clinical characteristics, and transmission patterns of a novel Mpox (Monkeypox) outbreak in eastern Democratic Republic of the Congo (DRC): *an observational, cross-sectional cohort study*

**DOI:** 10.1101/2024.03.05.24303395

**Authors:** Leandre Murhula Masirika, Jean Claude Udahemuka, Pacifique Ndishimye, Gustavo Sganzerla Martinez, Patricia Kelvin, Maliyamungu Bubala Nadine, Bilembo Kitwanda Steeven, Franklin Kumbana Mweshi, Léandre Mutimbwa Mambo, Bas B. Oude Munnink, Justin Bengehya Mbiribindi, Freddy Belesi Siangoli, Trudie Lang, Jean M. Malekani, Frank M. Aarestrup, Marion Koopmans, Leonard Schuele, Jean Pierre Musabvimana, Brigitte Umutoni, Ali Toloue, Benjamin Hewins, Mansi Dutt, Anuj Kumar, Alyson A. Kelvin, Jean-Paul Kabemba Lukusa, Christian Gortazar, David J Kelvin, Luis Flores

## Abstract

**Background:** In August 2023, an outbreak of mpox was reported in the eastern part, South Kivu Province, of Democratic Republic of the Congo. In this study, we aimed to investigate the origin of this outbreak and to assess how monkeypox virus spread among humans in the city of Kamituga.

**Methods:** We performed an observational cohort study by recruiting hospitalized patients with mpox-like symptoms. Furthermore, we compared structured, de-identified case report forms and interviews were conducted to determine the possible origins and modes of transmission of the mpox outbreak. We describe the clinical characteristics and epidemiology observed in reported infections.

**Findings:** During the study period (24 September 2023 to 29 January 2024), 164 patients were admitted to the Kamituga hospital, 51 individuals were enrolled in the study and interviewed, and 37 (73%) of 51 individuals received a molecularly confirmed mpox diagnosis. The median age for males was 24 years (IQR 18-30; range 14-36) and 19 years for females (IQR 17-21; range 1-59). The cohort was comprised of 47 (92%) of 51 individuals who identified as heterosexual, and two (4%) of 51 as bisexual, with 31 (61%) of 51 individuals sexually active with more than one partner within the last six months. The direct transmission routes are unknown; however, it is expected that the majority of infections were transmitted via occupational exposures. Out of the 51 individuals, 24 (47%) were professional sex workers (PSWs), while five (10%) were gold miners, 6 (12%) were students, and four (8%) were farmers; the remaining individual occupations were unknown. The most common symptoms associated with clinical mpox diagnosis were fever, which was described in 38 (75%) of 51 individuals, and rash, which was described in 45 (88%) of 51 individuals. Among those with a rash, 21 (41%) of 51 individuals experienced oral lesions, and 32 (63%) of 51 presented anogenital lesions. Mpox viral DNA was detected by qPCR from vaginal, penile, and oral swabs in 37 (73%) of 51 enrolled individuals. Two deaths were reported.

**Interpretation:** In this observational cohort study, mpox virus infection caused symptoms in a wide age range of participants with most cases presenting in sexually active individuals. Symptoms included fever, cough, lymphadenopathy, sore throat, chills, headache, back pain, muscle pain, vomiting, nausea, conjunctivitis, and rash (oral and anogenital). Heterosexual partners dominated human-to-human contact transmission suggesting that heterosexual close contact is the main form of transmission in this outbreak. Furthermore, Professional Sex Workers (PSWs) were the dominant occupation among infected individuals, indicating that PSWs and clients may be at higher risk for developing mpox virus infections.

## INTRODUCTION

Human mpox disease (formerly known as monkeypox) is a viral zoonotic disease caused by the monkeypox virus (MPXV). MPXV is a double-stranded DNA virus that belongs to the genus *Orthopoxvirus* within the *Poxviridae* family, subfamily *Chordopoxvirinae*. The clinical manifestations associated with mpox virus resemble those of smallpox -a disease which has been globally eradicated since 1977- and include rash (genital, anorectal, or mouth), fever, chills, fatigue, and sore throat (Alakunle et al., 2020; Dutt et al., 2023). MPXV was first isolated in 1958 during an outbreak at an animal research facility, where pox lesions were observed in macaque monkeys shipped to Copenhagen, Denmark, from Singapore (Brown et al., 2016, WHO). The first human case was reported in a 9-month-old boy in the Democratic Republic of Congo (DRC) in 1970. Since then, there have been sporadic mpox outbreaks with increasing frequency primarily in West and Central African countries, where the virus is considered to be naturally endemic (Moore et al, 2023). Before the 2022 multi-country mpox outbreak, the virus was divided into two distinct clades: (i) Clade I (formerly known as the Congo Basin or Central African clade) and Clade II (formerly known as the West African clade). Since August 2022, Clade II is further divided into two subclades, Clade IIa and Clade IIb (Schwartz et al., 2023). Viruses in Clade I and Clade II have a nucleotide sequence similarity of 99.4% (Schwartz et al., 2023; Chen et al., 2005). According to a study published in 2005, genomes of Clade I and Clade II are found to be highly conserved with minor differences at the sequence level (Likos et al., 2005; Americo et al., 2023). MPXV Clade I causes a more severe disease with a case fatality rate (CFR) of up to 10.6%, while MPXV Clade II is associated with milder symptoms and a lower CFR of roughly 1% (Shantier et al., 2022). MPXV contains a linear, double-stranded DNA genome with a length of roughly 200 kb, which encodes for ∼190 proteins. These proteins play important roles in DNA replication, transcription, and virion assembly (among others) as well as contributing to host viral immune system activation and evasion (Srivastava et al., 2023, Shantier et al., 2022).

Mpox is known to be more severe in individuals with comorbidities such as advanced Human Immunodeficiency Virus (HIV) infection (Mitjà et al., 2023; Saldana et al., 2023), uncontrolled diabetes mellitus (type one), and organ transplant (Riser et al., 2023). However, little is known about the disease outcome of mpox virus among patients with cancer (Demarco, 2022). Demographic risk factors also predispose certain groups to mpox virus infection. For example, the risk of contracting Clade IIb viral infection is increased for men who have sex with men (MSM) or for people who engage in sexual activity with infectious individuals (Amer et al., 2023). Mpox outbreaks have also been reported throughout the COVID-19 (coronavirus disease of 2019) pandemic. The impact of MPXV and SARS-CoV-2 virus and SARS-CoV-2 (severe acute respiratory syndrome coronavirus 2) co-infections remains to be fully elucidated due to sample size limitations (El-Qushayri et al., 2022). However, MPXV and SARS-CoV-2 co-infection was reported in one patient living with HIV (Nolasco et al., 2023).

There are two well-documented, classical routes of mpox virus transmission. The first is Primary zoonotic transmission, which originates in rodents and other small mammals and has been frequently associated with the Clade I viruses. Clade I mpox exhibits minimal human-to-human spread, resulting in a 10% rate of human mortality (Americo, Earl, and Moss, 2023). The animal reservoir of MPXV is not known with certainty, but small mammals (rope and sun squirrels), giant- pouched rats, and non-human primates (monkeys) are thought to maintain the virus in areas of West and Central Africa (Moore et al, 2023). However, non-sexual human-to-human transmission has also been documented with Clade I mpox virus (Nolen et al, 2016). Human-to-human transmission is associated with Clade IIb (the clade responsible for the 2022 global mpox outbreak) and linked to sexual contact with an infectious individual, resulting in genital and anorectal lesions that typically follow lesions at the site of infection.

In our observational cohort study, we propose a third route of MPXV transmission during the ongoing Clade I mpox outbreak in the city of Kamituga, South Kivu province (DRC). Here, we report cases of heterosexual Clade I mpox virus transmission and describe the characteristics of genital lesions among professional sex workers. Additionally, sustained transmission in Kamituga is occurring at the community level and being driven by a distinct Clade I mpox strain, possibly a novel subgroup, as confirmed with qPCR.

## METHODS

### Study design and participants

In this hospital-based mixed-methods prospective observational cohort study, we analyzed the demographic and clinical characteristics of all individuals with mpox virus infection symptoms admitted at the Kamituga Hospital between September 2023 and January 2024. We also discuss our findings compared with existing evidence. Written consent for the anonymized publication of images was individually sought from participants and documented. The ethical clearance to conduct this study was obtained from the Ethical Review Committee of the Catholic University of Bukavu (Number UCB/CIES/NC/022/2023). All study participants were introduced to the observational study and given the option to participate by providing informed consent or in the case that the participant was a minor, parental permission or assent was obtained. A mixed-methods study design was used, where the quantitative data were collected and analyzed first, and the qualitative data were collected second to elucidate the transmission cycles of mpox in the city of Kamituga. We confirm the IDs we used make the study participants unidentifiable.

### Study area

Kamituga is South Kivu’s largest gold mining city located in the territory of Mwenga, part of the South Kivu province. The South Kivu province borders in the north with the North Kivu province, south and west with the Maniema province, and south with the Tanganyika province. At the east, South Kivu borders Rwanda, Burundi, and Tanzania. The city has more than 260,000 inhabitants (based on the 2019 Kamituga Health Zone report), of which ∼16,000 are employed in the mining industry (Nkuba et al., 2017). The remaining residents depend directly or indirectly on ASGM (Artisanal and small-scale gold mining) for their livelihood.

### Procedures and sample collection

Routine data regarding age, gender, professional occupation, clinical presentation, concomitant presence of sexually transmitted infections (STIs), and comorbidities were collected from patient records (or hospital investigation forms) and entered into a secured, anonymized database. A questionnaire was also administered to collect information on sexual history, including sexual practices and number of sexual partners, as well as previous contacts, for each enrolled individual. Data were collected by trained field investigators using a pre-defined questionnaire based on Open Data Kit (ODK) software (an open-source data collection platform) installed on tablets with paper forms as backup. All data entered on the tablets were encrypted and only completed forms were transcribed and transmitted to the main application server.

Admission to the Kamituga Hospital was based upon clinical diagnosis of human mpox virus infection by hospital staff. A confirmed mpox virus case was defined as an individual with laboratory-detected infection. A case was listed as “suspect” if a patient had an acute illness with fever, intense headache, myalgia, and back pain, followed by one to three days of a progressively developing rash often starting on the face and spreading on the body. Finally, a case was listed as “probable” if it satisfied the clinical definitions of suspected cases and had an epidemiological link to a confirmed or probable case but was not laboratory-confirmed. The suspect cases were PCR tested in the Lwiro Laboratory (Centre de Recherche en Sciences Naturelles de Lwiro, CRSN- Lwiro). A skin lesion was defined as a single circumscribed area and included presentations comparable to papules, pustules, fluid-filled vesicles, or eschars. We defined a rash as a skin area, demarcated or diffusely distributed, with changes in skin texture or colour and inflammatory change.

### Statistical analysis

Descriptive statistics, including frequencies and proportions, were computed to gain a comprehensive understanding of the dataset’s characteristics. Exploratory Data Analysis (EDA) was applied in the epidemiological data, i.e., admissions and symptoms. The associations between hospitalization status and categorical variables (age groups, gender, and occupation) were explored, providing a nuanced understanding of potential relationships using Fisher’s exact test. Fisher’s exact test was chosen for its appropriateness with a relatively small sample size and situations involving sparse contingency tables. Fisher’s exact test statistics along with the corresponding p-values were used as an indicator of the statistical significance of the association between variables. The p-values were compared using a significance threshold of 0.05. Gender specific mixing patterns were analyzed by constructing a 2×2 contact matrix C with y_i j representing the total number of reported contacts between patients in gender category j and individuals in gender category i, where i = 1,2 and j = 1,2. Moreover, we performed chi-square tests to calculate the significance of a group (i.e., males and females) in answering “yes” or “no” to questions regarding their presented symptoms.

### DNA extraction and amplification

The DNA was manually extracted in a biosafety cabinet with a double-vaccinated scientist using the DNeasy Blood and Tissue kit (QIAGEN) following the manufacturer’s instructions. Briefly, samples were first treated with a lysis buffer (ATL) and then digested in proteinase K. AL buffer and alcohol were then added, and the mixture was collected into the DNeasy mini spin columns. After centrifugations and washing with buffer AW1 and AW2, the DNA was eluted with buffer AE, collected in Eppendorf DNA LoBind® tubes and quantified with a Qubit 4.0 fluorometer. The qPCR master mix was prepared with 20µl of Taqman polymerase, 2.7µl of the forward primers, 2.7µl of the reverse primers, 1.3µl of the probes, 3.3µl of nuclease-free water, and 10µl of DNA template, totaling 40µl. We used generic primers and probes (CDC) to detect the mpox viral DNA as recommended by the CDC for a qPCR assay. The qPCR assay was set at 50°C for two minutes followed by the activation of the polymerase at 95°C for 10 minutes. The amplification was performed at 95°C for 15 seconds and then dropped to 60°C for 30 seconds using the FAM fluorophore assignment and capturing for 45 cycles. The qPCR assay was performed on an X 960 real-time PCR machine (HealForce).

## RESULTS

A total One hundred and sixty-four mpox cases were listed as confirmed or suspected (Figure 1) as of January 2024. The highest number of admissions in a single week, i.e., 14, was reported in weeks 43, 48, and 52 in 2023.

**Figure 1.**
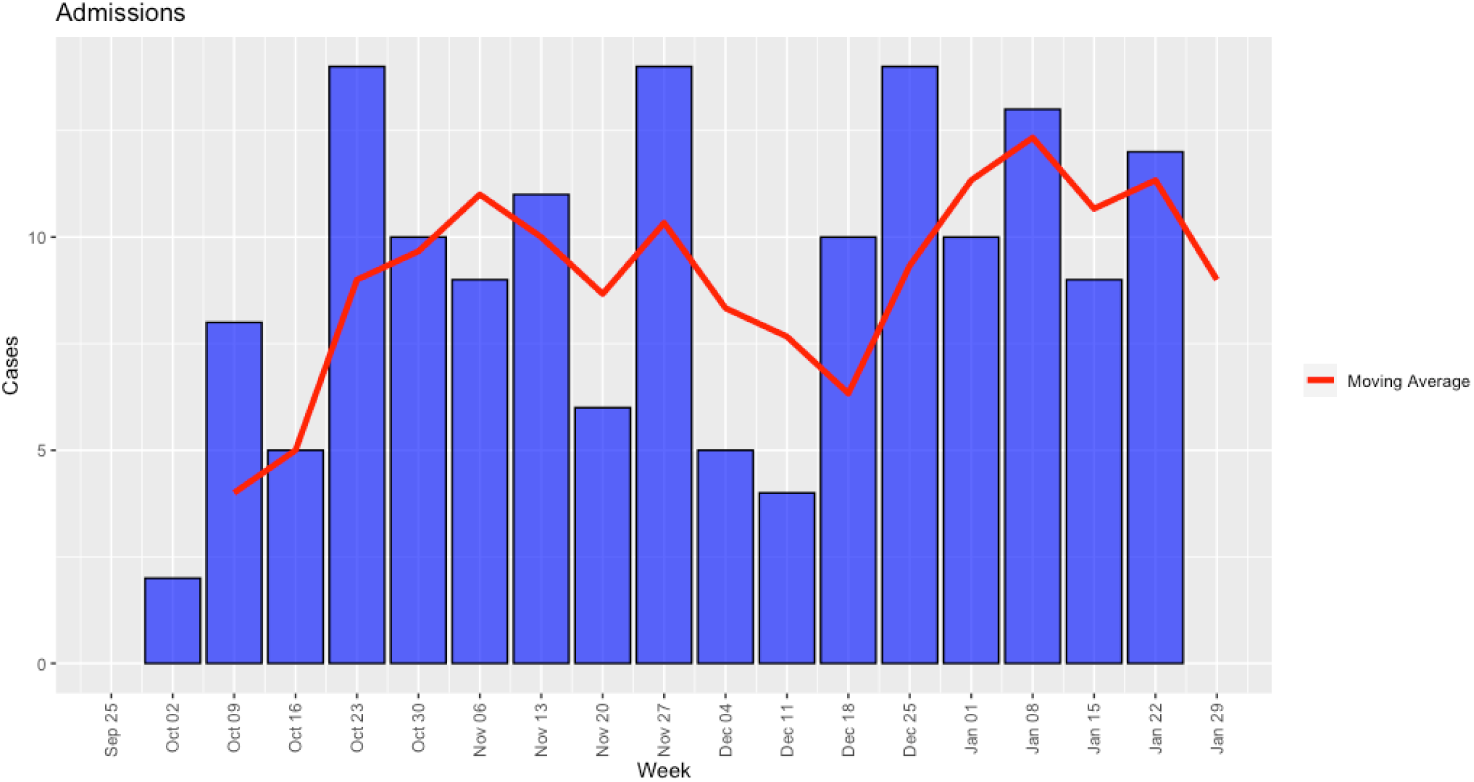
Epidemiological curve.

Of the total 164 patients admitted to the Kamituga hospital, 51 were enrolled in the study and were interviewed (Table 1). Of these 51 patients, 36 were PCR tested for viral load, 14 samples had a CT value less than 25, 5 patient samples had a CT value between 26 and 35, and 6 patient samples had a CT value greater than 36. The patients were also PCR tested in the surveillance program by the iNRB facility in Goma, North Kivu, DRC (only 2 of the 51 tested negative). CT values are pending for the remaining enrolled patients in the study. The median age of the female and male populations was 20 and 23, respectively. The sexual orientation of the patients is predominantly heterosexual (84.6% of the interviewed women and 100% of the interviewed men). Also, 61.5% of the interviewed women and 60% of the interviewed men reported having more than one sexual partner over the past six months. The profession most associated with female patients was found to be PSWs, encompassing 73% of the female population. In addition, five male individuals were also involved with the sex business in Kamituga. The single most predominant profession of the male respondents was gold mining (20%). No patients had previously received any mpox vaccination. Regarding the clinical symptoms of the confirmed/suspect cases, we report skin rash, fever, headache, and lymphadenopathy as the most common symptoms, having been reported by 88%, 74%, 72%, and 70%, respectively, of the entire cohort. We also report that two patients (one male and one female) reported being asymptomatic (Figure 2). The symptoms of skin rash, chills or sweats, genital lesions, headache, and muscle pain had significant *p* values, indicating there is an association between the groups and the symptoms.

**Figure 2.**
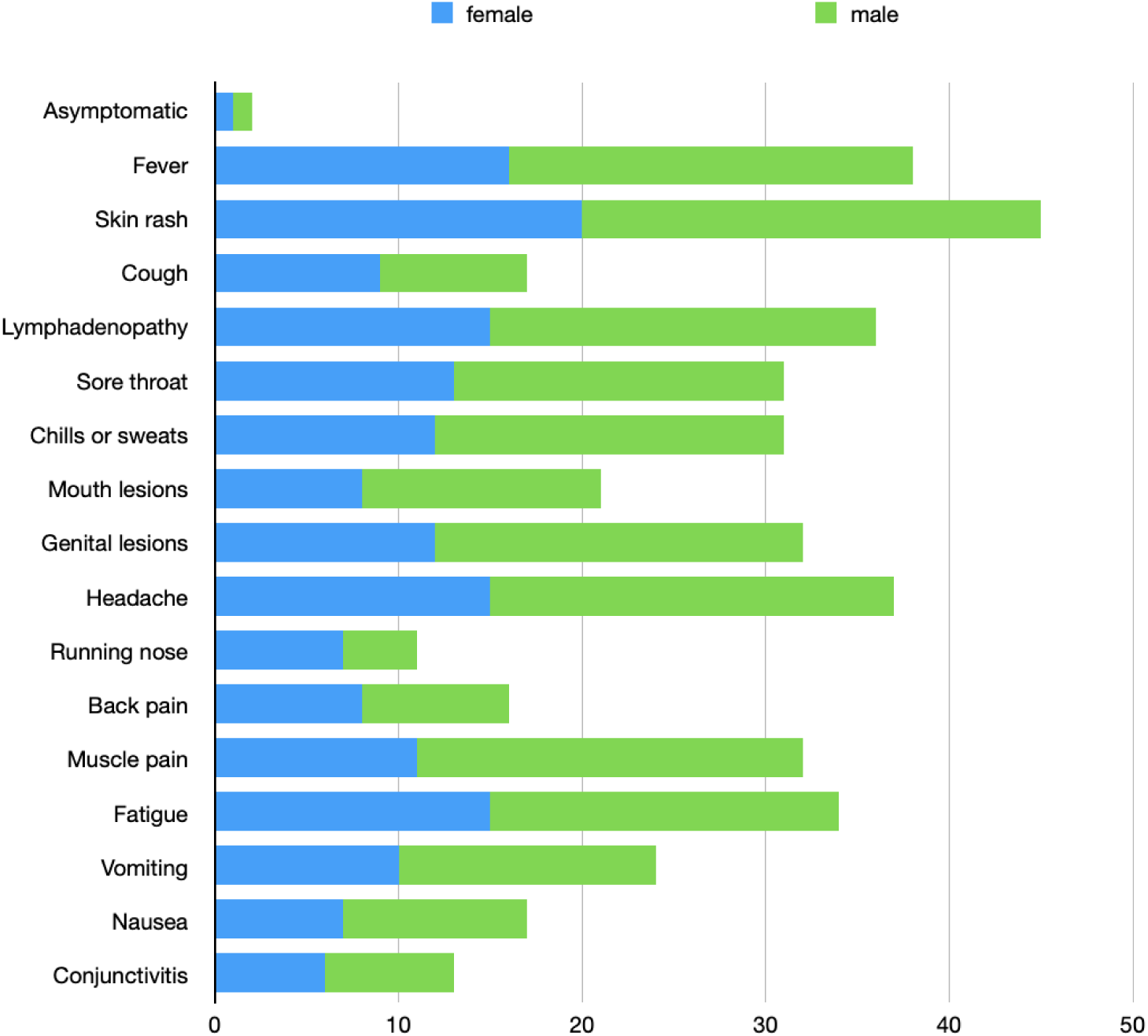
Symptoms.

**Table 1.**
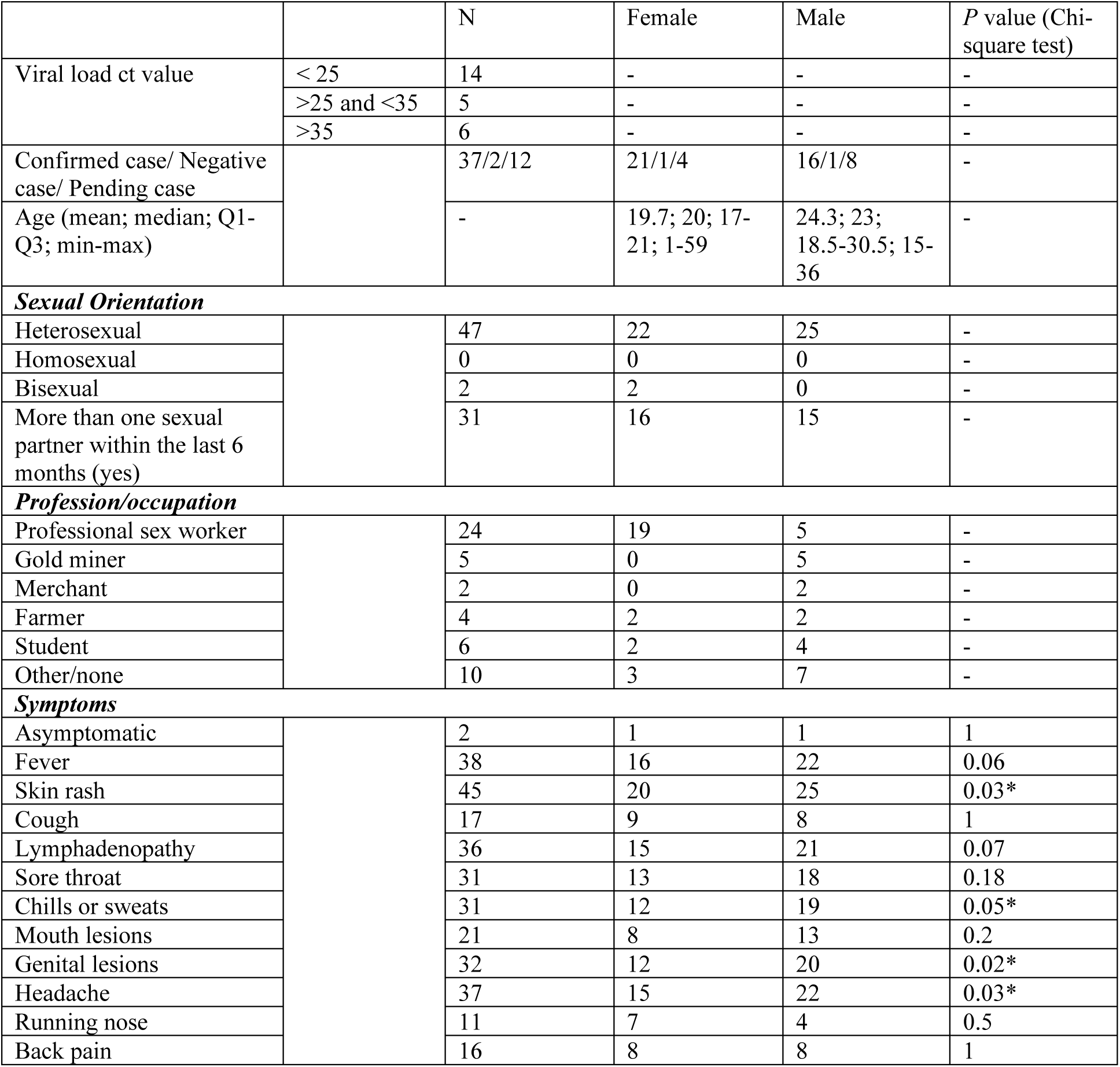
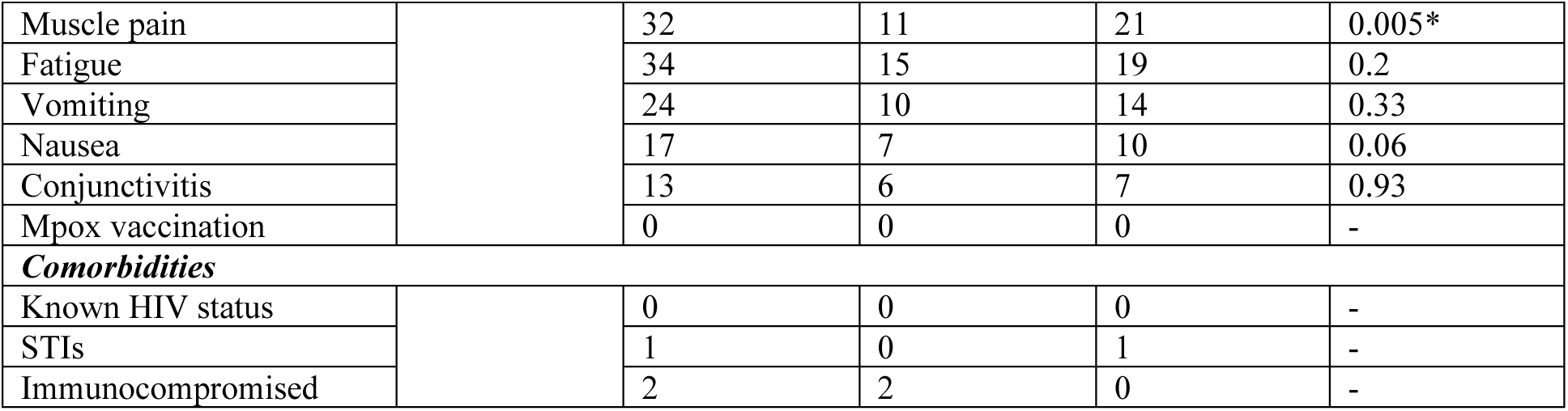
Characteristics of the cohort.

|  |  | N | Female | Male | P value (Chi-square test) |
| --- | --- | --- | --- | --- | --- |
| Viral load ct value | < 25 | 14 | - | - | - |
|  | >25 and <35 | 5 | - | - | - |
|  | >35 | 6 | - | - | - |
| Confirmed case/ Negative case/ Pending case |  | 37/2/12 | 21/1/4 | 16/1/8 | - |
| Age (mean; median; Q1-Q3; min-max) |  | - | 19.7; 20; 17-21; 1-59 | 24.3; 23; 18.5-30.5; 15-36 | - |
| ***Sexual Orientation*** |  |  |  |  |  |
| Heterosexual |  | 47 | 22 | 25 | - |
| Homosexual |  | 0 | 0 | 0 | - |
| Bisexual |  | 2 | 2 | 0 | - |
| More than one sexual partner within the last 6 months (yes) |  | 31 | 16 | 15 | - |
| ***Profession/occupation*** |  |  |  |  |  |
| Professional sex worker |  | 24 | 19 | 5 | - |
| Gold miner |  | 5 | 0 | 5 | - |
| Merchant |  | 2 | 0 | 2 | - |
| Farmer |  | 4 | 2 | 2 | - |
| Student |  | 6 | 2 | 4 | - |
| Other/none |  | 10 | 3 | 7 | - |
| ***Symptoms*** |  |  |  |  |  |
| Asymptomatic |  | 2 | 1 | 1 | 1 |
| Fever |  | 38 | 16 | 22 | 0.06 |
| Skin rash |  | 45 | 20 | 25 | 0.03* |
| Cough |  | 17 | 9 | 8 | 1 |
| Lymphadenopathy |  | 36 | 15 | 21 | 0.07 |
| Sore throat |  | 31 | 13 | 18 | 0.18 |
| Chills or sweats |  | 31 | 12 | 19 | 0.05* |
| Mouth lesions |  | 21 | 8 | 13 | 0.2 |
| Genital lesions |  | 32 | 12 | 20 | 0.02* |
| Headache |  | 37 | 15 | 22 | 0.03* |
| Running nose |  | 11 | 7 | 4 | 0.5 |
| Back pain |  | 16 | 8 | 8 | 1 |
| Muscle pain |  | 32 | 11 | 21 | 0.005* |
| Fatigue |  | 34 | 15 | 19 | 0.2 |
| Vomiting |  | 24 | 10 | 14 | 0.33 |
| Nausea |  | 17 | 7 | 10 | 0.06 |
| Conjunctivitis |  | 13 | 6 | 7 | 0.93 |
| Mpox vaccination |  | 0 | 0 | 0 | - |
| **Comorbidities** |  |  |  |  |  |
| Known HIV status |  | 0 | 0 | 0 | - |
| STIs |  | 1 | 0 | 1 | - |
| Immunocompromised |  | 2 | 2 | 0 | - |

We mapped the transmission of mpox virus within the community of Kamituga (Figure 3). The earliest documented case, Citizen (C) 1 initially had contact, of heterosexual nature, with citizens i.e., C2, who started showing Mpox symptoms in early October 2023, C3, who started showing Mpox symptoms in late September 2023, C4 who started showing Mpox symptoms in early October 2023, C5 who started showing Mpox symptoms in late September 2023, and c6, who started showing Mpox symptoms on early October 2023, all C1-5 were positively diagnosed with Mpox by PCR. In addition, C7, had contact with medical items used to treat C1, C7 was PCR positive for Mpox and started showing symptoms on early October 2023. These six initial contacts characterized the first link of the transmission chain. The second link of the transmission chain was characterized by C3 who had contact, of non-sexual nature, with C8, who started developing Mpox symptoms in mid-October 2023. C5 had contact, of heterosexual nature, with C9, who developed symptoms in late September 2023 and was PCR diagnosed with Mpox. C6 had contact of heterosexual nature with C10 who started developing symptoms in mid-October 2023 and was PCR diagnosed with Mpox. Finally, C7 had contact with C11, who started developing symptoms in early October 2023. These four contacts characterized the second link of the transmission chain. The third link of the transmission chain had C10, who had contact of heterosexual nature with C12, who started showing symptoms in late October 2023 and was PCR diagnosed with Mpox. Also, C11 had contact of non-heterosexual nature with C13 who started developing symptoms in late October 2023. Finally, C13 had contact with other 95 unconfirmed people and C1 had contact with other 25 unconfirmed people.

**Figure 3.**
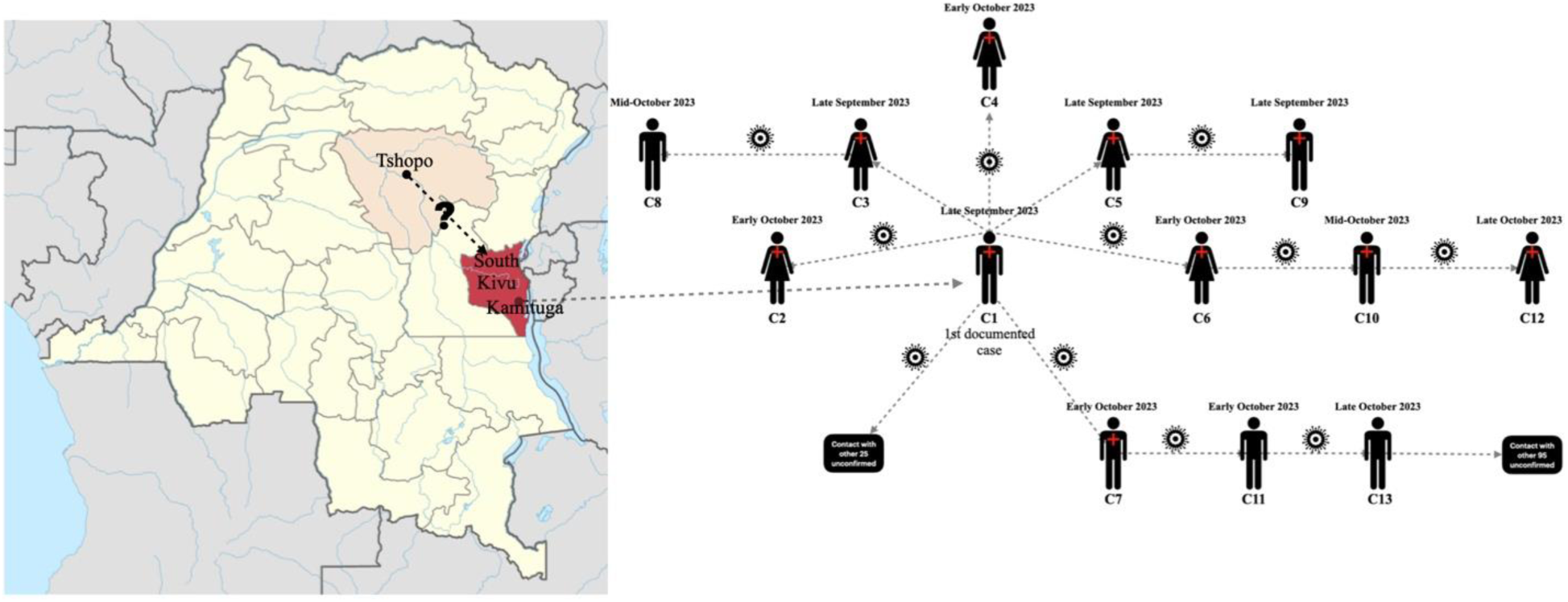
Suspected transmission cycles of mpox virus in the city of Kamituga.

We quantitatively tracked the contacts of the patients. Thirty individuals had contacts traced and were found to have had contact with 420 people (**Figure 4-A**). Most of the contacts occurred between males and females (**Figure 4-B**). We also report the nature of the contact forms (**Figure 4-C**) as being “contact with secreted bodily fluids of the infected patient, including blood, saliva, urine, or feces”, followed by “shared room” contact, in which individuals stayed in the same house and shared a room with infected individuals. The forms of contact “direct contact with the skin lesions”, which includes sexual contact, and “contact with wildlife”, were the least reported. Finally, we report on the relationships of individuals with people infected or those suspected of infection with mpox virus (**Figure 4-D**). The most common relationship was familial, followed by classmates and work colleagues.

**Figure 4.**
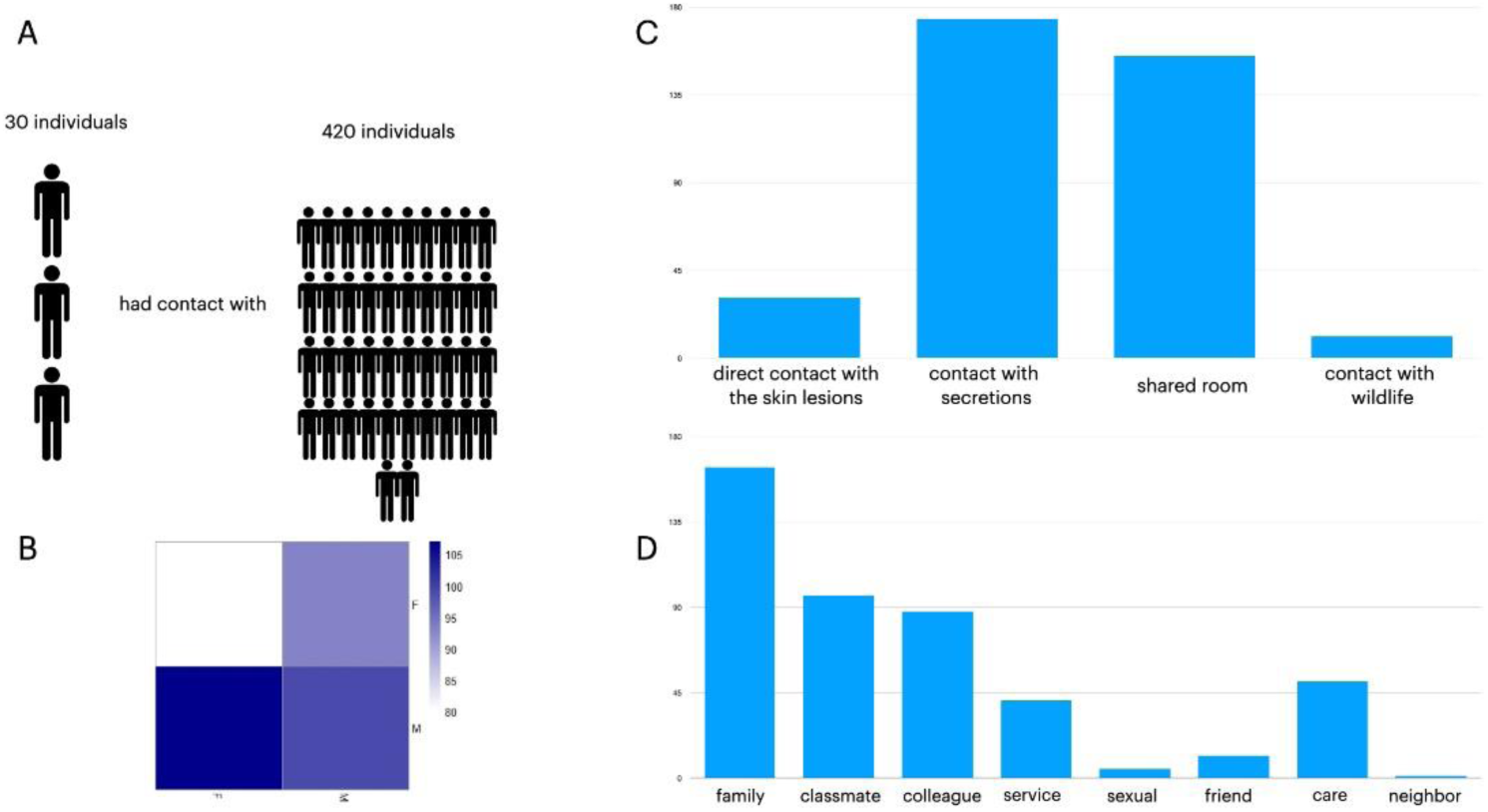
Contact tracing between individuals and people infected or suspected of infection by mpox virus.

## Discussion

In our observational cohort study, we describe an mpox Clade I virus outbreak whose initial spread was via heterosexual contact with infected individuals but was later transmitted among people who live with infected individuals or had other modes of direct or indirect contact. The early transmission cycles were characterized by heterosexual contact; later, the profile of the infected individuals included those with close contact such as intrafamilial, students, miners, farmers, caregivers, and merchants.

In 2022, a global outbreak of mpox (Clade IIb) was declared. During the 2022 outbreak, the primary demographic driver of infection was within the men who have sex with men (MSM) community, which reduced the possibility of spread to other demographic groups. Interestingly, few formal cases of Clade I were documented during the outbreak and prior to April 2023, no formal cases of sexual transmission of Clade I were reported. The first documented case of sexually transmitted mpox Clade I virus was in a male resident of Belgium, who had traveled to the DRC and had sexual encounters in the Kwango province (southwest) (Zebardast *et* al, 2023). Before 2023, the South Kivu province (Eastern DRC) had not previously reported cases of mpox (Makasa et al., 2021). In this study, our data suggests consistent mpox virus transmission over at least a 30- day window in Kamituga, a city located in Eastern DRC. We also observed a shift in the infected population, which transitioned from predominantly male to predominantly female.

Kamituga is known for its abundant fertile volcanic soil and numerous gold mines. Due to the concentration of gold mines, and thus gainful employment, Kamituga routinely welcomes individuals from neighboring territories and from other provinces within DRC. Individuals commonly travel to Kamituga for employment, especially since its nomination as the headquarters of La Compagnie Miniere de Grands Lacs Africains, a Belgian mining company, in 1923. Additionally, frequent cross-border travel occurs as individuals from neighboring countries specifically, Rwanda and Burundi, traveling to and from the city. Since the main activity and economic driver is gold mining, the sex work industry has served as a gainful occupation for females as PSWs and for males who act as managers of sex sites. Consequently, men and women residing in Kamituga live in overcrowded areas, characterized by poor hygienic conditions and a lack of contraceptives as well as basic necessities. Gold miners, sellers, and merchants often leave their families to work in Kamituga to provide a living wage for their families. Additionally, the bar managers leave Kamituga to recruit sex workers for their bars. Taken together, the socioeconomic characteristics of Kamituga and the surrounding region supports sexual and close contact virus transmission.

The Eastern Congo region has experienced armed conflict since the 1990s, which has drastically impaired health systems and basic individual access to healthcare services (Altare et al., 2021). Furthermore, the dire state of the roads connecting Kamituga to Bukavu (KivuTimes, 2023) makes it extremely difficult to transport equipment and supplies to the city. For example, a trip of 200 km can take up to two days during a health emergency, further exacerbating the situation. In our study cohort, the first four groups of people that had symptoms included PSWs, unemployed individuals, students, and gold miners. Except for the students, the other three groups follow a previously established link between mining sites and sexually transmitted diseases in Sub-Saharan Africa (reference). In Sub-Saharan Africa, it has been reported that HIV positivity likely doubles due to the likelihood of individuals having multiple sex partners, and condomless sex increases by 70% in areas with operating mines. Similarly, the relationship with high-risk sex partners increases by 30% (Magak, 2022, AIDSmap). Kamituga likely follows a similar trend, contributing to the high severity of symptoms and the longer period of clinical manifestations in mpox virus infected individuals. For the school students who displayed symptoms, these data coincide with other reports that have stated that in Africa the seroprevalence among children can be up to 40% (WHO, 2022) due to pupils having more physical and social contacts at school, especially during sports activities (Amzat et al., 2023).

During our investigation, we encountered challenges attributable to the absence of comprehensive diagnostic testing within the target demographic with respect to the identification and characterization of comorbidities. A more thorough picture of the immunocompromised population would have substantially augmented our understanding and interpretation regarding mpox viral evolution, which potentially exacerbates infection in individuals living with human immunodeficiency Virus (HIV).

In conclusion, we report on an ongoing outbreak of Clade I mpox in the Eastern DRC city of Kamituga, characterized by a previously undocumented model of heterosexual and community-level transmission. This outbreak has resulted in sustained transmission in Kamituga with over 200 cases identified since September 2023. We also identified a shift in the susceptible population of mpox, in which predominantly young women (median age 19) working as PSWs account for the majority of infected individuals during the initial cycles of transmission. Our data also indicates ongoing intrafamilial transmission of mpox virus in Kamituga with origins likely associated with the sex industry in the city.

## Acknowledgements

We greatly thank Wildlife Conservation Network (WCN) and Conservation Action Research Network (CARN) for the scholarship and research supports they awarded to the first author.

We would like to thank the Provincial Division of Health (DPS) of South-Kivu and Kamituga Health Zone (KHZ) for their collaboration during the study.

Finally, we greatly thank the EU Horizon 2020 grants VEO (874735) and the Global Health EDCTP3 Joint Undertaking (Global Health EDCTP3) program under grant agreement No. 101103059 (GREATLIFE) for the technical collaboration during this outbreak.

## Author Approval

All authors approved the final version of the manuscript.

## Data availability

Clinical data is available upon reasonable request to the corresponding author.

